# Rare coding variants in five DNA damage repair genes associate with timing of natural menopause

**DOI:** 10.1101/2021.04.18.21255506

**Authors:** Lucas D. Ward, Margaret M. Parker, Aimee M. Deaton, Ho-Chou Tu, Alexander O. Flynn-Carroll, Gregory Hinkle, Paul Nioi

**Affiliations:** Alnylam Pharmaceuticals, Cambridge, MA 02142, USA

## Abstract

The age of menopause is associated with fertility and disease risk, and its genetic control is of great interest. We used whole-exome sequences from 119,992 women in the UK Biobank to test for associations between rare damaging variants and age at natural menopause. Rare damaging variants in three genes significantly associated with menopause: *CHEK2* (p = 6.2 × 10^−51^) and *DCLRE1A* (p = 1.2 × 10^−12^) with later menopause and *TOP3A* (p = 8.8 × 10^−8^) with earlier menopause. Two additional genes were suggestive: *RAD54L* (p = 2.3 × 10^−6^) with later menopause and *HROB* (p = 2.7 × 10^−6^) with earlier menopause. In a follow-up analysis of repeated questionnaires in women who were initially pre-menopausal, *CHEK2, TOP3A*, and *RAD54L* genotype associated with subsequent menopause. Consistent with previous GWAS, all five genes are involved in the DNA-damage repair pathway. Phenome-wide scans across 363,977 men and women revealed that in addition to known associations with cancers and blood cell counts, rare variants in *CHEK2* also associated with increased risk of uterine fibroids, polycystic ovary syndrome, and prostate hypertrophy; these associations are not shared with higher-penetrance breast cancer genes. Causal mediation analysis suggests that approximately 8% of the breast cancer risk conferred by *CHEK2* pathogenic variants after menopause is mediated through delayed menopause.

## Introduction

The age at natural menopause (ANM) varies widely between women and understanding the biology of menopause timing is important because early menopause is associated with risk of cardiovascular disease and osteoporosis, and late menopause is associated with risk of breast cancer^1^. ANM is also strongly related to fertility, because natural fertility ends on average ten years before menopause^2^.

Genetic and environmental factors both correlate with ANM. Lower socioeconomic status, fewer live births, not using oral contraceptives, and smoking have been consistently associated with earlier ANM^1^. Twin studies and family studies established that ANM is a highly heritable trait^3; 4^. Population-based genome-wide association studies (GWAS) have identified many common polymorphisms associating with ANM^5-11^. These ANM-associated common variants are enriched at loci harboring genes involved in the DNA repair and replication checkpoint process such as *BRCA1* [MIM: 113705], *MCM8* [MIM: 608187], *CHEK2* [MIM: 604373], and *HELB* [MIM: 614539]. A recent Mendelian randomization (MR) analysis found an association between common ANM-associated variants and breast cancer risk, but not conversely between common breast cancer-associated variants and ANM. This observation supports a causal relationship between variation in lifetime endogenous estrogen exposure (resulting from variation in the duration between menarche and menopause) and risk of breast cancer^11^.

GWAS are limited in their ability to identify causal genes because the majority of associated common haplotypes contain no coding variants, and the closest gene to a noncoding variant is not reliably the causal gene in the absence of functional evidence. Indeed, the variants associated with ANM near tumor suppressor genes *BRCA1* and *CHEK2* have not been the same pathogenic variants implicated in breast cancer. To better identify causal genes for ANM by discovering rare coding variants strongly associated with ANM, we used exome sequencing data from 119,992 women in the UK Biobank. After identifying genes where aggregated protein-truncating variants associated with ANM, we then performed phenome-wide analysis of quantitative traits and disease diagnoses in carriers of these variants across 363,977 exome-sequenced individuals in the UK Biobank.

## Methods

### UK Biobank study

The UK Biobank consists of approximately 500,000 volunteer participants, who were aged 40-69 when recruited between 2006 and 2010^12; 13^. Both array genotyping and whole exome sequencing has been performed on the majority of these participants. Data from genotyping, sequencing, questionnaires, primary care data, hospitalization data, cancer registry data, and death registry data were obtained through application number 26041.

### Variant calling and definition

The source of genetic data for the main analysis was exome sequencing data. DNA from whole blood was extracted and sequenced by the Regeneron Genetics Center (RGC) using protocols described elsewhere^14^. Of the variants called by RGC, additional quality control filters were applied: Hardy-Weinberg equilibrium (among the White subpopulation) p-value less than 10^−10^ and rate of missing calls across individuals less than 2%. Variants were then annotated using ENSEMBL Variant Effect Predictor (VEP) v95^15^, using the LOFTEE plug-in to additionally identify high-confidence predicted protein-truncating variants (PTVs, also known as predicted loss-of-function, pLOF)^16^. Variants were also annotated with the Whole Genome Sequence Annotator (WGSA)^17^ to add Combined Annotation Dependent Depletion (CADD) scores^18^ to predict deleteriousness of missense variants. For single-variant analyses, a minor allele frequency filter was imposed such that only variants present in at least ten individuals with phenotype data were retained. For rare variant burden analyses, variants were defined as rare if their minor allele frequency was under 1%. Variants were aggregated in each protein-coding gene as follows: PTV variants were defined as variants with their most severe consequence from VEP as “stop gained”, “splice donor”, “splice acceptor”, or “frameshift,” and their confidence from LOFTEE as “HC” (high confidence). Damaging missense variants were defined as variants with their most severe consequence from VEP as “missense” and a CADD score of 25 or greater. Genes were defined as implemented by ENSEMBL v95 and further filtered to retain only “genes with protein product” as currently defined by HGNC (accessed January 28, 2021)^19^.

A supplementary source of genetic data, used only for single-variant tests, was obtained from array genotyping. Genotypes were called through chip typing and imputation as described previously^13^. Variants were filtered so that imputation quality score was greater than 0.8, missingness across individuals was less than 2%, and minor allele frequency was at least ten carriers with data for the phenotype being analyzed.

### Participant definition for overall analyses

An initial round of quality control was performed by RGC which removed subjects with evidence of contamination, discrepancies between chromosomal and reported sex, and high discordance between sequencing and genotyping array data. Genetic relatedness was calculated using the PRIMUS algorithm^20^ and an “unrelated” population was defined by removing all first- and second-degree relatives, as well as some third-degree relatives. Principal components analysis (PCA) was used to define the White population as follows: among individuals in the unrelated set who identified as White in the self-reported ethnicity question (Field 21000), PCA was performed using high-quality common variants using the eigenstrat algorithm^21^; variants for this initial round of PCA were filtered for missingness across individuals < 2%, minor allele frequency > 1%, excluding regions of long-range LD^22^, and independence (pairwise LD r^2^ < 0.1). Principal components (PCs) were then projected onto the related individuals who were held out from the unrelated set, and all individuals greater than three standard deviations away from the mean of PCs 1-6 were removed as ancestry outliers. A final PC estimation was performed in eigenstrat on the remaining individuals, using the unrelated subjects for PC determination and projecting the related individuals onto these final PCs. This resulted in a set of 363,977 White unrelated individuals (398,574 when including related individuals for the follow-up SAIGE-GENE analyses that used them) with exome sequencing data available.

### Participant and phenotype definition for menopause analysis

For menopause analyses, an initial set of 216,554 unrelated White women with exome-sequencing data was considered. Self-reported data from the touchscreen questions administered at the initial assessment center visit were used as an initial filter. Women were only retained in downstream analyses if they answered that they had not yet had menopause, or that they had had menopause and knew the age at which it had occurred. Women were excluded if their answer to the menopause question was “not sure – had a hysterectomy” or “not sure – other reason.” Women were further asked wither they had ever had a bilateral oophorectomy or a hysterectomy and were excluded if they answered that they did not know. If they reported that they had had either procedure, they were excluded if they were not able to report the age at which they had had the procedure. Women were asked if they had used hormone-replacement therapy (HRT); women were excluded if they did not know or did not answer the question, and those who reported having used HRT were excluded if they were not able to report the age at which they had first used HRT. Women were excluded if the age they reported having an oophorectomy was earlier or the same age as the age they reported menopause; if the age they reported having a hysterectomy was earlier or the same age as the age they reported menopause; and if the age they reported first using HRT was younger than the age they reported menopause. Premenopausal women were excluded if they had ever used HRT or had a bilateral oophorectomy or hysterectomy.

Additional data about potentially menopause-inducing operations was then obtained from inpatient hospital diagnoses and operation codes. Hospital diagnoses for radiotherapy, chemotherapy for cancer or chemotherapy not otherwise specified, and follow-up care for those procedures, were obtained corresponding to ICD10 codes Z51.0, Z55.1, Z55.2, Z08.1, Z08.2, Z09.1, and Z09.2. Operations for chemotherapy, radiotherapy, hysterectomy, or bilateral oophorectomy were obtained corresponding to OPCS4 codes X35.2, X37.3, X38.4, X70, X71, X72, X73, X74, X65, X67, Y90.2, Y91, Y92, Q07, Q08, and Q22. The earliest possible calendar year of menopause was derived by adding the age at menopause to the year of birth. The earliest calendar year for any of these procedures for a woman was defined as the year of the procedure. Postmenopausal women were excluded if they had any of these procedures in the same year or earlier year than the earlies possible calendar year of menopause, and premenopausal women were excluded if they had had any of the procedures.

An additional 1,893 women were excluded from the main analysis because they had an age of menopause less than 40, and an additional 249 women were excluded because they reported menopause after 60 or were premenopausal and over 60 when interviewed. These women were used for subsequent replication analysis of extreme phenotypes. These exclusions resulted in a set of 119,992 women for the main analysis (132,377 women for the confirmatory SAIGE-Gene analyses that included related individuals.)

A more restrictive subset of menopause data was then constructed as a subset of the main analysis set (“neoplasm- and surgery-free”) which did not rely on the relative timing of potentially menopause-inducing operations and which excluded anyone with any neoplasm preceding the time of interview. Women were excluded if they reported having had bilateral oophorectomy, hysterectomy, or doctor-diagnosed cancer at the time of their initial assessment, regardless of the age at which these had occurred, and if they had any cancer or neoplasm diagnosis in the national cancer registries in the year of the interview or earlier. Women were also excluded based on procedures found in ICD10 hospital diagnoses and OPCS4 operation codes in a calendar year the same as or earlier than the interview, using the ICD10 and OPCS4 codes listed previously and also adding the following: ICD10 code Z40.0 (prophylactic surgery for risk factors related to malignant neoplasms) and OPCS codes corresponding to all of chapter Q (operations on the upper female reproductive tract), except for codes purely relating to diagnostic examination: Q18, Q39, Q50, or Q55. This “neoplasm- and surgery-free” subset consisted of 92,387 women.

### Time-to-event analysis

The main discovery analysis consisted of a time-to-event analysis of the association between carrier status of rare variants aggregated per gene and menopause age. For each variant set, a Cox proportional hazards model was constructed using the survival R package^23^. A right-censored survival object was created where the status indicator was zero for women who were premenopausal and one for women who were post-menopausal, and follow-up time was the age at interview for premenopausal (censored) women and age at menopause for postmenopausal women. Ties were handled using the default method (Efron approximation). With this survival object as the dependent variable, the independent variable in each model was the presence (coded as one) or absence (coded as zero) of at least one alternate allele from the variant set in each individual; multiple variants and zygosity were not considered for the burden test. Covariates were year of birth (to account for secular trends in menopause age^24^) and the first twelve genetic PCs (to account for associations arising purely from population stratification.)

Time-to-event analysis was also performed for individual exome and array-typed variants; these tests were performed as above, except the individual variable was the genotype of the individual variant, coded as zero (homozygous reference), one (heterozygous), or two (homozygous alternate.) When both exome and array data were available for a variant, the exome genotype data was used; when only array data was available for a variant, the association was only calculated for the subset of exome-sequenced individuals who also had array data available.

To determine the extent to which linkage disequilibrium was driving multiple associations seen in regional patterns of single-variant association results at the *CHEK2* and *RAD54L* loci, associations in the region were tested again including the genotypes of top association signals as covariates in the regression (rs34001746 at *CHEK2* and rs12142240 and rs12073998 at *RAD54L*.)

### Linear and logistic regression analysis

Secondary burden analyses were performed for menopause timing: menopause status at interview, as a case-control trait, was tested using logistic regression with age at interview, year of birth, and the first twelve genetic PCs as covariates; and age of menopause among postmenopausal women, as a quantitative trait, was tested using linear regression with year of birth and the first twelve genetic PCs as covariates. These regressions were performed using the glm function in R.

### Generalized mixed model analysis

Because of known issues with high Type 1 error rate when performing association tests between rare variants and rare diagnoses, and because of potential confounding by population stratification even when excluding related individuals, SAIGE-GENE^25^ was used for confirmation of some associations. SAIGE-GENE uses a generalized mixed model taking subject relatedness into account and uses a saddle point approximation to estimate p-values. SAIGE-GENE was performed across the entire White population, including related individuals, and a genetic relatedness matrix (GRM) was calculated using a set of 100,000 LD-pruned variants selected from across the allele frequency spectrum.

### PheWAS

For the variant sets significantly associated with menopause timing, phenome-wide association studies were performed between carrier status of variants in each set and 3,457 diagnosis codes and 99 quantitative traits. Diagnosis codes were obtained from a combination of inpatient hospital diagnoses (Field 41270), causes of death (Field 40001 and 40002), the national cancer registries (Field 40006), and general practitioner (GP) clinical event records which were available for a subset of individuals (Field 42040). Quantitative traits were selected from a variety of phenotypes available from UK Biobank fields, including anthropomorphic measurements, smoking behavior, blood and urine biochemistry, etc. and were inverse rank normalized using the RNOmni R package^26^. Quantitative traits and cancer registry diagnoses were downloaded from the UK Biobank Data Showcase on March 17, 2020. Because GP clinical events, inpatient diagnoses, and death registry were available in more detail or in more recent updates than the March 2020 Data Showcase download, these data were supplemented with data downloaded as separate tables: data for GP clinical records were downloaded on September 30, 2019, data from the death registry was downloaded on June 12, 2020, and data from hospital diagnoses was downloaded on July 15, 2020. Regressions were performed using linear or logistic regression as appropriate using the glm function in R, with age, sex, country of recruitment (England, Scotland, or Wales), availability of GP data, and the first 12 PCs as covariates.

### Mediation analysis

Mediation analysis was performed using the R package mediation^27^. Data from the cancer registry was used to limit the analysis to only test breast cancer occurring after 60 as cases and women who were breast cancer free and over 60 as controls. The analysis was also limited to women who reported an ANM between ages 40 and 60 at the initial interview, to ensure for simplicity of the model that breast cancer occurred after menopause.

## Results

### Gene-level associations with ANM

Among UK Biobank participants with exome sequencing data currently available, we identified a subset of unrelated White women for genetic analysis who had not experienced any treatments that would preclude natural menopause. Of these, 78,316 reported being postmenopausal and reported an age at menopause, while 41,676 reported being pre-menopausal at their initial interview (**Figure S1**). We defined two sets of rare exome variants to aggregate for each gene: one set consisting only of protein-truncating variants (PTVs), also known as predicted loss-of-function (pLOF) variants, and a second set including both PTVs and rare missense variants bioinformatically predicted to be deleterious. We analyzed variant sets in genes that had at least ten carriers (12,467 genes with enough PTV carriers and 16,078 genes with enough PTV and missense carriers). We performed three association tests for each gene: a burden test using current menopause status (pre- or post-) as a case-control trait in a logistic regression model, a burden test using ANM among post-menopausal women as a quantitative trait in a linear regression model, and a time-to-event (TTE) analysis using both groups jointly in a Cox proportional hazards model (**Table S1**; QQ plots and λ_GC_ calculations, **Figure S2**). In the TTE analyses, aggregated variants in three genes surpassed a conservative threshold Bonferroni corrected for the total number of genes and variant sets tested of p < 1.8 × 10^−6^: *CHEK2* (both variant definitions) and *DCLRE1A* [MIM: 609682] (both variant definitions) with later menopause, and *TOP3A* [MIM: 601243] (PTVs and missense combined) with earlier menopause. At a more lenient threshold correcting only for the number of genes tested for each variant set definition (p < 4.0 × 10^−6^ for PTVs only and 3.1 × 10^−6^ for PTVs and missense), two additional genes were associated: *RAD54L* [MIM: 603615] (PTVs and missense combined) with later menopause and *HROB* [MIM: 618611] (PTVs only) with earlier menopause (**Figure 1**). All five genes were separately associated with both ANM among postmenopausal women and menopause status at the time of the interview among all women (**Table 1**).

**Table 1.** Results for seven variant sets significantly associated with menopause timing. Three tests are shown for each variant set: (1) Time-to-event (TTE) analysis using Cox regression; resulting hazard ratio (HR) greater than one means carriers experienced earlier menopause and less than one means carriers experienced later menopause; number of events analyzed is N carriers post menopause; (2) Quantitative trait analysis of age of natural menopause (ANM) among post-menopausal women, using linear regression; resulting statistic is the modeled association of carrier status with ANM; (3) Case-control analysis of menopause status at interview (pre-vs post-menopausal), using logistic regression; resulting statistic is the odds ratio (OR), where values greater than one mean carriers were more likely to be post-menopausal at time of interview, and values less than one mean carriers were more likely to be pre-menopausal at time of interview.

| gene | variant set | N carriers total | N carriers post-menopause | Model | Phenotype | statistic | 95% CI | p value |
| --- | --- | --- | --- | --- | --- | --- | --- | --- |
| *CHEK2* | PTV and damaging missense | 1698 | 993 | Cox<br>linear<br>logistic | TTE<br>ANM<br>meno. status | HR = 0.62<br>1.46 yrs<br>OR = 0.37 | [0.58, 0.66]<br>[1.22, 1.71]<br>[0.30, 0.46] | 6.22e-51<br>3.37e-32<br>2.36e-20 |
| *CHEK2* | PTV only | 765 | 431 | Cox<br>linear<br>logistic | TTE<br>ANM<br>meno. status | HR = 0.52<br>2.22 yrs<br>OR = 0.26 | [0.47, 0.57]<br>[1.86, 2.59]<br>[0.19, 0.36] | 2.18e-41<br>1.80e-32<br>9.74e-17 |
| *DCLRE1A* | PTV and damaging missense | 4178 | 2681 | Cox<br>linear<br>logistic | TTE<br>ANM<br>meno. status | HR = 0.89<br>0.24 yrs<br>OR = 0.72 | [0.86, 0.93]<br>[0.09, 0.39]<br>[0.63, 0.82] | 5.84e-09<br>1.49e-03<br>1.67e-06 |
| *DCLRE1A* | PTV only | 1213 | 769 | Cox<br>linear<br>logistic | TTE<br>ANM<br>meno. status | HR = 0.77<br>0.61 yrs<br>OR = 0.53 | [0.72, 0.83]<br>[0.34, 0.89]<br>[0.42, 0.69] | 1.25e-12<br>1.28e-05<br>9.75e-07 |
| *TOP3A* | PTV and damaging missense | 2704 | 1764 | Cox<br>linear<br>logistic | TTE<br>ANM<br>meno. status | HR = 1.14<br>-0.37 yrs<br>OR = 1.27 | [1.09, 1.19]<br>[-0.55, -0.18]<br>[1.08, 1.50] | 8.88e-08<br>9.58e-05<br>4.23e-03 |
| *RAD54L* | PTV and damaging missense | 1911 | 1223 | Cox<br>linear<br>logistic | TTE<br>ANM<br>meno. status | HR = 0.87<br>0.35 yrs<br>OR = 0.71 | [0.82, 0.92]<br>[0.13, 0.57]<br>[0.58, 0.86] | 2.27e-06<br>1.81e-03<br>5.25e-04 |
| *HROB* | PTV only | 86 | 66 | Cox<br>linear<br>logistic | TTE<br>ANM<br>meno. status | HR = 1.78<br>-2.26 yrs<br>OR = 3.94 | [1.40, 2.27]<br>[-3.20, -1.33]<br>[1.40, 11.10] | 2.73e-06<br>2.17e-06<br>9.41e-03 |

**Figure 1.**
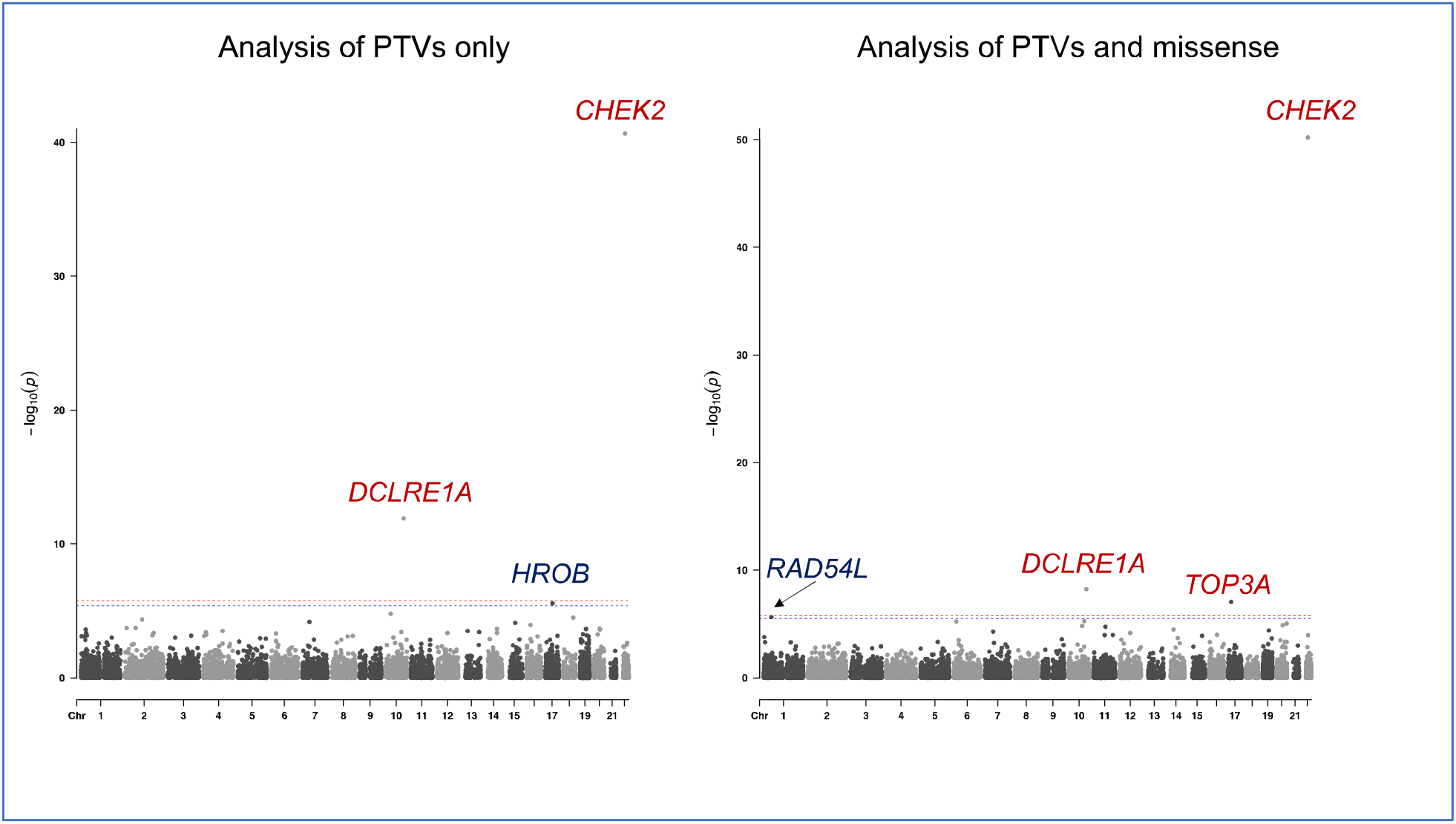
Manhattan plot of genome-wide scan of gene-level tests of menopause timing. P-value is from TTE analysis using a Cox proportional hazards model.

**Figure 2.**
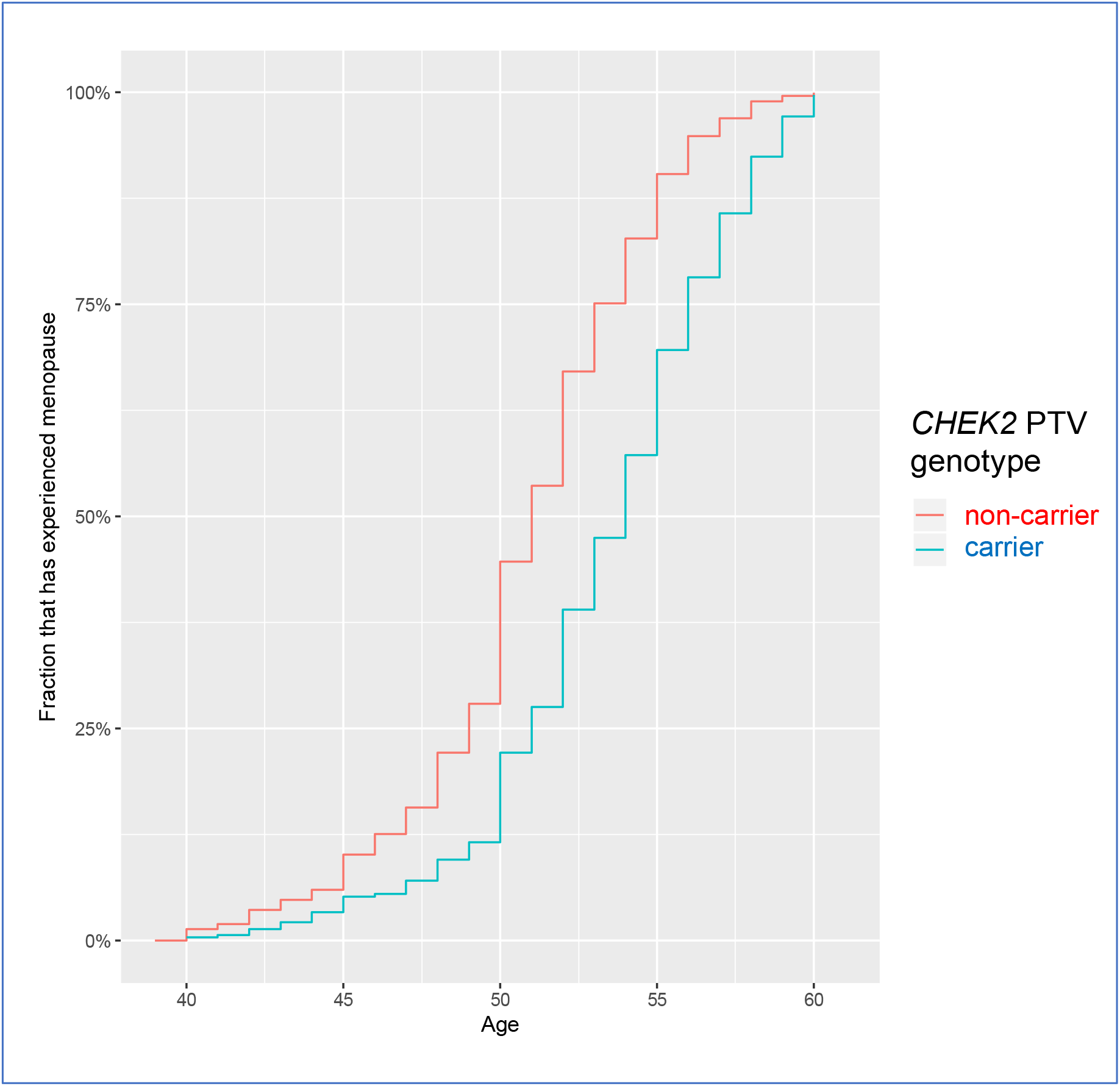
Cumulative incidence of menopause among CHEK2 PTV carriers and non-carriers between the ages of 40 and 60.

### Confirmatory analyses

To account for potential inaccuracy in recalling the timing of hysterectomy and oophorectomy relative to menopause in the interview data, which could have resulted in some residual surgically-induced menopause events in our analysis, we created a more conservative set of ANM values that excluded women with any self-reported or registry-recorded cancer or gynecological operations before the date of the questionnaire. All five genes remained associated with menopause in this subset of women (**Table S1**).

Although our discovery analysis excluded closely related individuals, cryptic relatedness cannot be excluded as a source of population stratification. To account for relatedness, we repeated the quantitative trait and case-control analyses using the SAIGE-GENE algorithm^25^, which accounts for genetic relatedness; because relatedness is explicitly modeled, we performed this analysis in an expanded set of 132,377 women which included related individuals. All of these associations remained significant by SAIGE-GENE (**Table S2**).

We sought additional evidence outside of the discovery dataset for these associations in two ways. First, we performed a TTE analysis in a subset of 4,701 women who were premenopausal at the initial interview and who participated in a follow-up interview between 2-14 years later, 3,630 of whom were subsequently postmenopausal and had a known ANM at the time of their most recent follow-up interview. For all seven variant sets associated in the discovery analysis, the 95% confidence interval in the follow-up analysis was consistent with that of the discovery analysis. For *CHEK2, RAD54L*, and *TOP3A*, rare variants associated menopause timing in the follow-up interview at p < 0.05 (**Table S3**; **Figure S3**). Second, we looked at two extreme phenotypes we had excluded from the discovery analysis: 1,893 women with primary ovarian insufficiency (POI), defined as menopause before the age of forty, and 249 women with menopause after the age of sixty. At a threshold of p < 0.05, we detected a depletion of *CHEK2* and *DCLRE1A* carriers and an enrichment of *TOP3A* carriers in women with POI, and an enrichment of *CHEK2* carriers in women with menopause after sixty (**Table S4**).

### Association of individual rare and common variants with ANM

To more thoroughly evaluate genetic mechanisms of association with menopause timing at these five loci, we tested all individual coding variants contributing to these associations with at least ten carriers (**Table S5**). The most significant association in *CHEK2* was rs555607708, a frameshift variant (p.Thr367MetfsTer15; commonly known as c.1100del; p = 1.6 × 10^−27^) well-studied as a pathogenic variant in breast and other cancers, and the second most significant association in *CHEK2* was rs587780174, another pathogenic frameshift variant (p.Ser422ValfsTer15; commonly known as c.1263del; p = 1.7 × 10^−9^); the strongest missense association was with rs28909982 (p.Arg117Gly; p = 1.3 × 10^−4^). The most significant association in *DCLRE1A* was nonsense (premature stop) variant rs41292634 (p.Arg138Ter; p = 7.5 × 10^−11^) and the strongest missense association in *DCLRE1A* was missense variant rs11196530 (p.Ile859Phe; p = 1.7 × 10^−3^). The most significant association in *TOP3A* was missense variant rs34001746 (p.Leu584Arg; p = 1.6 × 10^−10^). The most significant association in *RAD54L* was missense variant rs28363218 (p.Arg202Cys; p = 7.1 × 10^−5^) and the most significant association in *HROB* was nonsense rs774881553-T (p.Gln341Ter; p = 2.7 × 10^−3^). We also re-ran the tests of aggregated variants with a leave-one-out approach to test the extent to which these individual variants contributed to the gene-level associations. For all genes except *TOP3A*, the gene-level association remained significant when holding out the strongest single-variant associations; for *TOP3A*, missense rs34001746 appeared to explain the entire association (p = 0.50 for all other missense and PTVs.)

We then broadened our analysis to 30,571 array-genotyped or exome-sequenced variants within each of the five gene bodies and 500 kilobases flanking, focusing on associations that would be considered significant in a GWAS (p < 5 × 10^−8^; **Table S6**; **Figures S4-S8**). No additional variants at *DCLRE1A* or *HROB* were associated with menopause timing. At *TOP3A*, four noncoding variants in addition to missense rs34001746 were associated, had the same MAF as rs34001746 (0.7%), and were in strong linkage disequilibrium (**Figure S6**). At both *RAD54L* and *CHEK2*, many common and rare variants were significantly associated, consistent with a previous GWAS of ANM by the ReproGen consortium^11^ which identified noncoding associations at both of these loci and hypothesized *RAD54L* and *CHEK2* as the causal genes. We explored the independence of the associations with the multiple variants at these loci by building TTE models with genotypes of multiple variants as dependent variables.

At *RAD54L*, conditional analysis suggested that the common GWAS variant rs12142240 (reported by ReproGen) and rs12073998 (the strongest common-variant association we find, which is in high linkage disequilibrium with rs12142240) were in high linkage disequilibrium with all of the other genome-wide significant associations at the locus, and rare missense rs28363218 remained nominally significant at p = 2.6 × 10^−3^ suggesting its independence (**Table S7**). At *CHEK2*, conditioning on the genotype of the common GWAS variant rs34001746 reduced the number of additional genome-wide significant SNP associations from 34 to 7, while improving the significance of the association with rare frameshift rs587780174 (**Table S8**). In a TTE model incorporating both rs34001746 and rs587780174, both remain highly significant (p = 2.9 × 10^−10^ and p = 9.2 × 10^−28^ respectively), suggesting that the association identified by GWAS is independent of the predominant rare variant signal at the *CHEK2* locus.

### Other associations with menopause timing-associated genes

To better understand the full spectrum of consequences of rare coding variants in these five genes, we performed phenome-wide association studies (PheWAS) for association with 3,457 diagnosis codes and 99 quantitative traits. While *DLCRE1A, RAD54L, TOP3A*, and *HROB* had no associations reaching phenome-wide significance (p < 2.0 × 10^−6^), *CHEK2* associated with many diagnoses and traits (**Table S9; Table 2**).

**Table 2.** Selected associations of *CHEK2* damaging variants with diagnoses and quantitative traits. Associations shown are for *CHEK2* PTVs and missense variants aggregated; all associations are also significant when considering *CHEK2* PTVs only. Cases observed are counted among the carriers; the expected value of case carriers is based on the prevalence of the disease and carrier frequency.

| phenotype | p | Effect | N |
| --- | --- | --- | --- |
| Platelet crit | 4.95e-39 | beta = 0.18 SD [0.15, 0.20] | 4939 carriers measured |
| Leukocyte count | 4.60e-29 | beta = 0.16 SD [0.13, 0.19] | 4939 carriers measured |
| C50: Malignant neoplasm of breast | 2.58e-27 | OR = 1.87 [1.67, 2.10] | 351 cases / 199 expected |
| Mean corpuscular hemoglobin | 2.79e-21 | beta = -0.13 SD [-0.16, -0.10] | 4939 carriers measured |
| Lymphocyte count | 1.89e-20 | beta = 0.13 SD [0.10, 0.16] | 4932 carriers measured |
| Neutrophil count | 1.41e-19 | beta = 0.13 SD [0.10, 0.16] | 4932 carriers measured |
| D05: Carcinoma in situ of breast | 1.47e-14 | OR = 2.18 [1.79, 2.65] | 105 cases / 49 expected |
| C61: Malignant neoplasm of prostate | 2.82e-12 | OR = 1.67 [1.44, 1.92] | 222 cases / 144 expected |
| D25: Leiomyoma of uterus | 4.34e-10 | OR = 1.49 [1.32, 1.69] | 278 cases / 190 expected |
| Erythrocyte count | 1.40e-08 | beta = 0.07 SD [0.04, 0.09] | 4939 carriers measured |
| E28.2: Polycystic ovarian syndrome | 1.47e-08 | OR = 3.05 [2.07, 4.49] | 28 cases / 9 expected |
| N40: Hyperplasia of prostate | 2.47e-08 | OR = 1.40 [1.24, 1.57] | 361 cases / 281 expected |
| C92: Myeloid leukemia | 5.03e-07 | OR = 2.74 [1.85, 4.06] | 26 cases / 10 expected |
| L82: Seborrhic keratosis | 6.44e-07 | OR = 1.31 [1.18, 1.45] | 419 cases / 329 expected |

Besides breast cancer and related diagnoses, rare damaging variants in *CHEK2* were also associated with prostate cancer, which has been previously reported^28^. *CHEK2* variants also associated with myeloid leukemia, consistent with reports that *CHEK2* coding variants associate with myelodysplastic syndrome^29^. We also found associations with benign neoplasms: carcinoma in situ of the breast and leiomyoma of the uterus (uterine fibroids.) In addition to these neoplasms, we also detected association between *CHEK2* variants and diagnosis of polycystic ovary syndrome (PCOS), prostate hyperplasia, and seborrheic keratosis. Because highly imbalanced case-control ratios can lead to false positives in association tests, we confirmed that these associations remained significant when using the SAIGE-GENE algorithm, which corrects for these potential false positives and also accounts for relatedness **(Table S10)**. Among quantitative traits, *CHEK2* variants associate with many hematological parameters, most strongly with increased lymphocyte count and increased platelet crit. Multiple hematological associations with *CHEK2* were noted in a recent study of 49,960 exome sequenced UK Biobank participants, which the authors noted could be secondary to cancer treatment^14^. To test whether these associations were shared with other breast cancer genes, we performed PheWAS on rare damaging variants in four other moderate-to high-penetrance breast cancer genes (*BRCA2* [MIM: 600185], *BRCA1, PALB2* [MIM: 610355], and *ATM* [MIM: 607585])^30^ and found that the hematological associations and disease associations, with the exception of carcinoma in situ of the breast and prostate cancer, were specific to *CHEK2* (**Table 3**; **Table S11**).

**Table 3.** Testing *CHEK2*-associated phenotypes with four other breast cancer genes. For uniformity of comparison, for all genes, PTVs only were aggregated and tested. Genes were listed if the association p < 0.05/56. A plus (+) indicates that the 95% confidence interval of the estimated effect is greater and non-overlapping with the corresponding interval for *CHEK2*.

| phenotype | associated breast cancer genes |
| --- | --- |
| Platelet crit | *CHEK2* |
| Leukocyte count | *CHEK2* |
| C50: Malignant neoplasm of breast | *BRCA2+*, *BRCA1+*, *PALB2+*, *CHEK2*, *ATM* |
| Mean corpuscular hemoglobin | *CHEK2* |
| Lymphocyte count | *CHEK2* |
| Neutrophil count | *CHEK2* |
| D05: Carcinoma in situ of breast | *BRCA2*, *PALB2*, *CHEK2*, *ATM* |
| C61: Malignant neoplasm of prostate | *BRCA2*, *CHEK2*, *ATM* |
| D25: Leiomyoma of uterus | *CHEK2* |
| Erythrocyte count | *CHEK2* |
| E28.2: Polycystic ovarian syndrome | *CHEK2* |
| N40: Hyperplasia of prostate | *CHEK2* |
| C92: Myeloid leukemia | *CHEK2* |
| L82: Seborrheic keratosis | *CHEK2* |

We further tested the non-neoplasm associations in two sub-analyses: an analysis of men who have never been diagnosed with a neoplasm, and an analysis of women who have never been diagnosed with a neoplasm which also conditioned on menopause status as a covariate. All associations except seborrheic keratosis were replicated in these secondary analyses, suggesting that these associations were not secondary to cancer treatment, diagnosed neoplasms, or menopause status, and that the hematological associations were not sex-specific (**Table S12**).

Because of the association between estrogen exposure and menopause timing, we also tested all of the ANM-associated variant sets against estradiol levels, limiting this analysis to 32,385 premenopausal women with measurements available. No association was detected (**Table S13**).

### Mediation analysis of *CHEK2* effects on menopause and breast cancer

The association of *CHEK2* variants with both breast cancer and delayed menopause raised the question of the extent to which this pleiotropy was mediated by the well-established association between delayed menopause and breast cancer. To distinguish between biological and mediated pleiotropy, we performed causal mediation analysis. For simplicity of the model, we used only the subset of women who were postmenopausal at the time of recruitment and who reported an ANM. To further ensure that breast cancer followed menopause and that genetic effects on ANM did not confound the association between the age covariate and follow-up time, we only considered breast cancer cases vs. breast cancer-free controls after the age of 60. In this subset, we detected significant associations of *CHEK2* genotype with ANM, *CHEK2* genotype with breast cancer, and ANM with breast cancer (while controlling for *CHEK2* genotype), suggesting both a direct effect and indirect effect (via menopause delay) of *CHEK2* genotype on breast cancer. Using PTVs only, which have the largest estimated effect on breast cancer, we tested the significance of the unstandardized indirect effect using 1,000 bootstrapped samples and estimated the bootstrapped indirect effect as 7.8% of the total effect (95% CI: 4.5% to 14.9%; p < 2 × 10^−16^; **Figure 3**). The proportion was similar when considering both PTVs and missense variants (7.1% mediated; 95% CI: 4.1% to 15.5%; p < 2 × 10^−16^; **Figure S9**).

**Figure 3.**
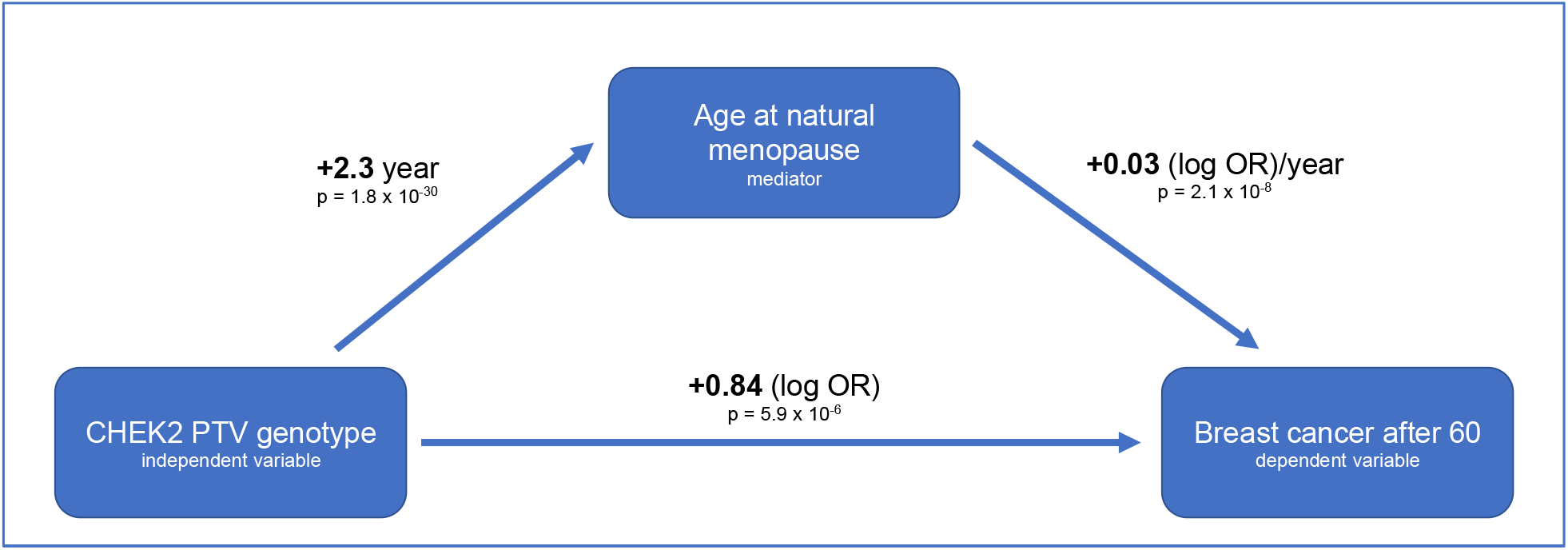
Mediation analysis of the proportion of CHEK2 PTV’s breast cancer effect mediated by delaying ANM. The bottom arrow shows the modeled total effect of CHEK2 genotype increasing risk of breast cancer after 60 (from a logistic regression model). The upper left arrow shows the modeled amount by which CHEK2 delays ANM (using linear regression among postmenopausal women). The upper right arrow shows the modeled effect of delaying ANM by one year on risk of breast cancer after 60 (using logistic regression).

## Discussion

### Genetic architecture of menopause timing

Deciphering the genetic control of menopause is important for understanding the causal relationship between menopause and associated disease risks. Association studies have revealed many loci influencing menopause timing, but the majority of association signals have consisted of common noncoding variants. Additionally, these studies have only looked at ANM in postmenopausal women, discarding potentially useful information from the dichotomous trait of whether women are post-or pre-menopausal at a given age. To increase power and improve causal gene discovery, we performed time-to-event analysis including both pre- and post-menopausal women and used a large cohort of exome sequences from the UK Biobank to interrogate rare damaging variants. The largest published GWAS of menopause timing, from the ReproGen consortium, identified 44 associated loci, 29 of which included at least one gene involved in DNA damage repair^11^. However common coding variants identified causal genes unambiguously at only five loci: *EXO1* [MIM: 606063], *REV3L* [MIM: 602776], *HELB, BRCA1*, and *MCM8*, all of which encode genes involved in DNA damage repair. Other associated haplotypes harbored coding variants in *PRIM1* [MIM: 176635], *FANCI* [MIM: 611360], *GSPT1* [MIM: 139259], *NLRP11* [MIM: 609664], and *SLCO4A1* [MIM: 612436], but there were additional potential causal genes at these loci; of these, two are involved in DNA damage repair. The largest GWAS of menopause timing in non-European ancestry women, performed in Japanese women, identified additional associated loci with coding variants in *GNRH1* [MIM: 152760], *ZCCHC2*, and *ZNF518A* [MIM: 617733]^*7*^. Our analysis identified rare coding variant associations with menopause timing in five additional genes: *CHEK2, TOP3A, DCLRE1A, RAD54L*, and *HROB*. All five are involved with various aspects of DNA damage repair.

The strong link between DNA damage repair genes and menopause timing is not surprising given the role of DNA damage repair and surveillance at every stage of the development of oocytes^11; 31^. The size of the initial oocyte pool at birth, along with the rate of atresia of oocytes through life, influences the age at which the oocyte pool is depleted to a number low enough to trigger amenorrhea (approximately 1,000 remaining oocytes). The meiosis that occurs in oocytes necessitates programmed double-stranded breaks (DSBs) which must be repaired through the homologous recombination pathway; oocytes that do not properly repair DSBs after this first phase of meiosis undergo apoptosis. Oocytes rest for decades in an arrested meiotic state until ovulation, during which they accumulate additional DNA damage from exogenous insults; the mouse homolog of *CHEK2* has been shown to cause oocytes to undergo apoptosis when they have experienced damage from ionizing radiation^31; 32^.

Mendelian randomization analysis of menopause-delaying alleles supports a causal role of these variants in breast cancer risk, mediated through prolonged exposure to endogenous estrogen^11^. The plausibility of this causal mechanism is strongly supported by the fact that hormone-replacement therapy is a risk factor for breast cancer^33^. Although common variants at the *CHEK2* and *BRCA1* loci have been previously associated with ANM, the rare coding variants that associate with breast cancer at these loci have not been previously reported to associate with ANM at genome-wide significance. A recent preprint identified common-variant associations at *CHEK2* and *BRCA2* with ANM and followed up these associations by testing for association between *CHEK2* and *BRCA2* pathogenic variants and menopause timing among 9,619 exome-sequenced women; they found associations with later menopause (p = 1 × 10^−5^) and earlier menopause (p = 0.03), respectively^34^. Although we find an association between *BRCA2* PTVs and earlier menopause in our primary analysis (p = 3.2 × 10^−4^), this association is largely abrogated when excluding any women with history of neoplasms or gynecological surgery (p = 0.08), suggesting the primary association could be confounded by prophylactic or therapeutic surgery in *BRCA2* carriers. The association we observe between *CHEK2* rare damaging variants and later menopause timing is strong and genome-wide significant even at the level of individual variants and is seen in additional data not used in the discovery analysis. Through causal mediation analysis, we show for the first time that while the predominant effect of *CHEK2* pathogenic variants is directly on breast cancer risk, that risk would be slightly less if it were not for their menopause-delaying effect.

### Novel *CHEK2* biology

In addition to discovering associations between rare damaging variants in *CHEK2* with menopause timing, and replicating known associations with breast cancer^30^, prostate cancer^28^, and myeloid leukemia^29^, we find associations with many hematological measurements such as increased leukocyte counts and platelet crit, and associations with several diagnoses: polycystic ovary syndrome (PCOS), uterine fibroids, prostate hyperplasia, and seborrheic keratosis. These hematological associations were first reported in an earlier analysis of the UK Biobank exome data and were hypothesized to be secondary to cancer treatment^14^; however, we find that they exist in individuals with no diagnosed neoplasm. They are unlikely to be secondary to menopause timing, as they persist in both men and women, and are seen among women even when correcting for menopause status at the time of the blood assay. It is more likely, therefore, that these hematological changes arise through the same mechanisms that predispose to myelodysplastic syndrome in *CHEK2* mutation carriers^29^. After *TP53, CHEK2* was identified as a second causal gene for Li-Fraumeni syndrome (MIM: 609265), which involves risk for many cancers^35; 36^. A common feature of the non-cancer diseases we find associated with *CHEK2* is that they all involve benign hyperproliferation of tissue. Although *CHEK2* acts as a brake on proliferation in response to DNA damage, it may serve as a more general negative regulator of mitosis in somatic cells even in the absence of DNA damage.

### Genetics and molecular biology of *DCLRE1A, TOP3A, RAD54L*, and *HROB*

*DCLRE1A* (DNA cross-link repair protein 1a, also known as the homolog of *S. cerevisiae* SNM1) encodes a protein that repairs inter-strand crosslinks^37^; it has not been implicated in any Mendelian syndromes or GWAS of complex traits. Nonsense variants such as rs41292634 and missense variants such as rs11196530 both contribute to the ANM association we identify with *DCLRE1A*, which was not detected in prior GWAS of ANM. While rs41292634 has been reported as a candidate SNP in BRCA1- and BRCA2-negative cancer families^38^, *DCLRE1A* has not been detected in larger exome-sequencing studies. We also observe a depletion of *DCLRE1A* carriers in POI cases not included in our discovery analysis.

*TOP3A* (DNA topoisomerase III alpha) encodes a protein critical to DNA synthesis by dissolving double Holliday junctions after homologous recombination-mediated repair of double-stranded DNA breaks^39; 40^. It has been implicated in a recessive developmental disorder (MGRISCE2, related to Bloom syndrome [MIM: 618097]) characterized by short stature and microcephaly. We find an ANM association at *TOP3A* that is entirely explained by a rare missense variant rs34001746, reported as likely benign in ClinVar; this association is the strongest rare variant association we detect with earlier menopause. Consistent with the direction we detect, *TOP3A* rs34001746 carriers are enriched in cases of POI not included in our discovery analysis, and carriers who were premenopausal at their initial interview were more likely than non-carriers to be postmenopausal at followup interviews.

The remaining loci, *RAD54L* and *HROB*, are genome-wide significant in the individual genome-wide scans but do not surpass a strict Bonferroni-corrected threshold for having performed two genome-wide scans. Whether such a conservative threshold is justified is debatable given that variants were shared between the two scans, and they were therefore far from independent tests. We chose to follow up and report on these results in light of prior biological evidence, while noting that replication in a different cohort would be critical before considering these associations high confidence.

*RAD54L* (Rad54-like, named after its *S. cerevisiae* homolog) encodes a protein involved in homologous recombination that binds tightly to Holliday junctions^41^. Although *RAD54L* has been proposed as a gene altered in the germline in breast cancer^42; 43^, it has not been associated in modern exome-wide studies of breast cancer^30^, and consistent with this we do not observe an association with breast cancer in the UK Biobank. The ReproGen GWAS of ANM identified a noncoding association at the *RAD54L* locus; we identify an independent rare variant association of later ANM with missense rs28363218, which has not been reported as pathogenic to ClinVar. We find that the association of *RAD54L* with later ANM is confirmed by follow-up interviews of initially premenopausal women.

*HROB* (homologous recombination factor with OB-fold, recently renamed from *C17orf53*) encodes a protein that is involved in homologous recombination by recruiting the MCM8-MCM9 helicase after DNA damage^44^. HROB-deficient mice are infertile, with ovaries lacking follicles in females and defective sperm production in males. Missense variants in *MCM8* have been previously identified in GWAS of ANM and nonsense variants in *MCM9* have been implicated in POI (MIM: 616185)^45^. While *HROB* PTVs are exceedingly rare in our study (only 86 carriers), precluding power to replicate in our subgroup analyses, evidence from mouse phenotypes and the human genetics of the epistatic *MCM8* and *MCM9* genes make the association with *HROB* highly biologically plausible.

### Limitations

The problems inherent in studies of on self-reported recollection of age at menopause have been previously reported^1; 46^. The accuracy of recall of age at menopause is known to decrease with duration between menopause and the interview. A bias towards ages divisible by five (40, 45, 50) is clear from the distribution of ages and suggests low accuracy of individual reports of ANM. Oral contraceptive use may mask the onset of menopause but is so widespread that excluding oral contraceptive users was infeasible for our study.

Except for the most common pathogenic *CHEK2* variants which have been characterized in the context of breast cancer, the other variants we identify as associating with ANM remain to be tested functionally to validate bioinformatic predictions. Experimental work is needed to verify that these variants indeed disrupt protein function (in the case of missense variants) or result in nonsense-mediated decay of the principal message (in the case of PTVs).

While we have attempted to use follow-up interviews and extreme phenotypes to obtain additional evidence to confirm the initial discoveries, a true replication would involve study of a separate cohort that has both menopause information and rare variant information from exome or whole-genome sequencing. Such independent replication will be critical to confirming the weaker evidence seen for associations with *RAD54L* and *HROB*.

### Conclusions

Our study of rare coding variants confirms findings from previous GWAS highlighting a key role of DNA damage repair proteins in genetic determination of menopause timing. In addition to identifying coding variant associations at the *CHEK2* and *RAD54L* loci previously identified by GWAS, we identify novel associations at *TOP3A, DCLRE1A*, and *HROB*, and confirm that menopause timing is the sole phenome-wide significant association for rare variants in these genes. *CHEK2* also appears to be highly pleiotropic beyond its known role in breast cancer and other cancer syndromes, affecting hematological traits as well as conferring risk for benign disorders involving hyperproliferation of tissue such as PCOS, prostate hyperplasia, and seborrheic keratosis.

## Data Availability

The data used in this study were obtained from the UKBB through application 26041. All phenotypic data and array genotypes are accessible through application to UK Biobank. Currently, exome sequencing data for ∼200,000 participants is available; the remainder of the exome data is scheduled for public release in 2021.

## Ethics Statement

Approval was received to use these data from the UK Biobank under application number 26041. The UK Biobank resource is an approved Research Tissue Bank and is registered with the Human Tissue Authority, which means that researchers who wish to use it do not need to seek separate ethics approval (unless re-contact with participants is required). Ethics oversight for the UK Biobank is provided by an Ethics and Governance Council which obtained written informed consent from all participants for use of the data in health-related research.

## Acknowledgements

This research has been conducted using the UK Biobank resource, application number 26041. We thank the UK Biobank participants for their donations to this resource. Data management and analytics were performed using the REVEAL/SciDB translational analytics platform from Paradigm4.

## Supplemental Figures

**Figure S1.**
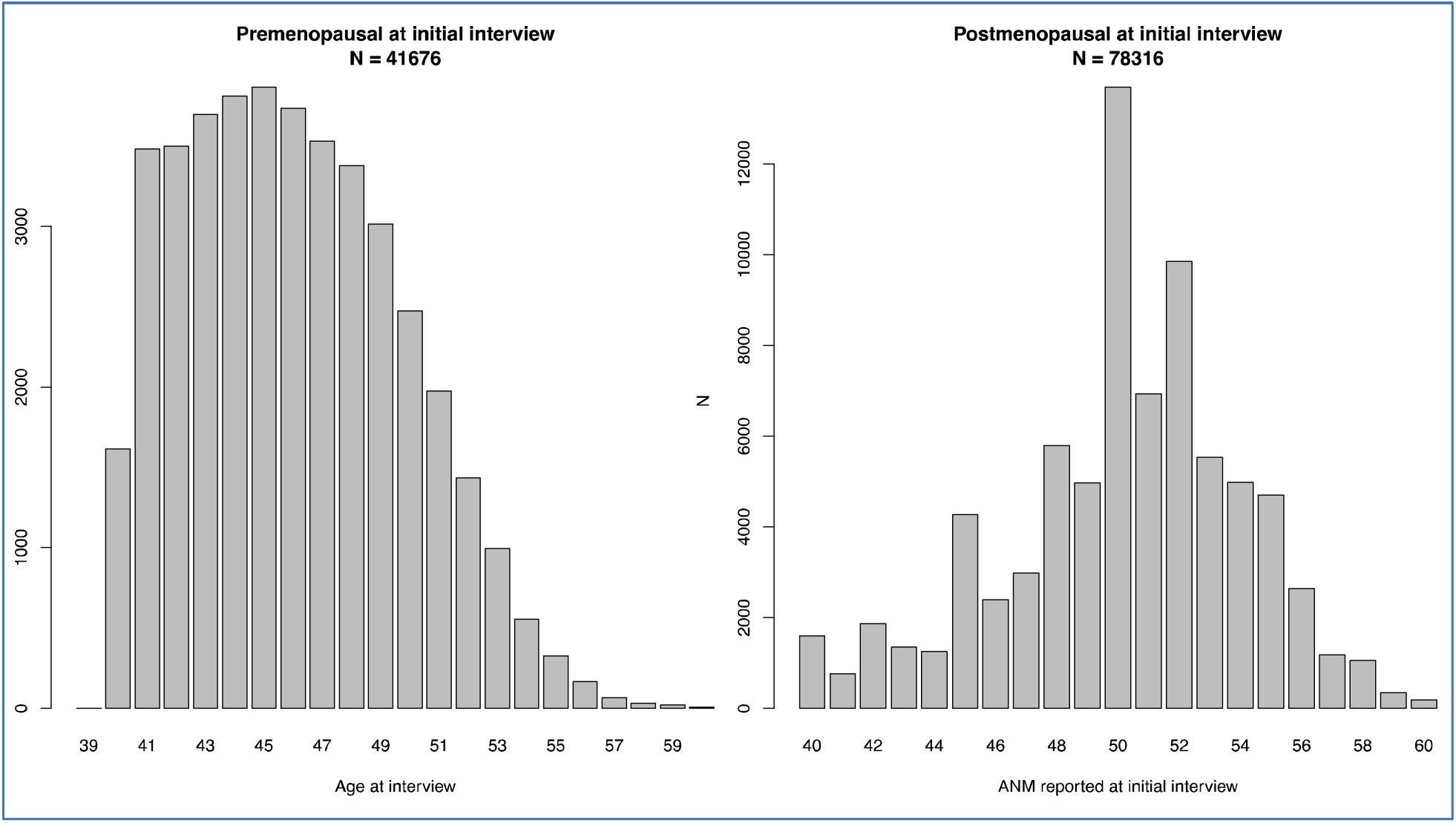
Summary of phenotype data used for association analysis. (L) Premenopausal women and age at the interview at which they report being premenopausal. (R) Postmenopausal women and the age at which they reported experiencing natural menopause.

**Figure S2.**
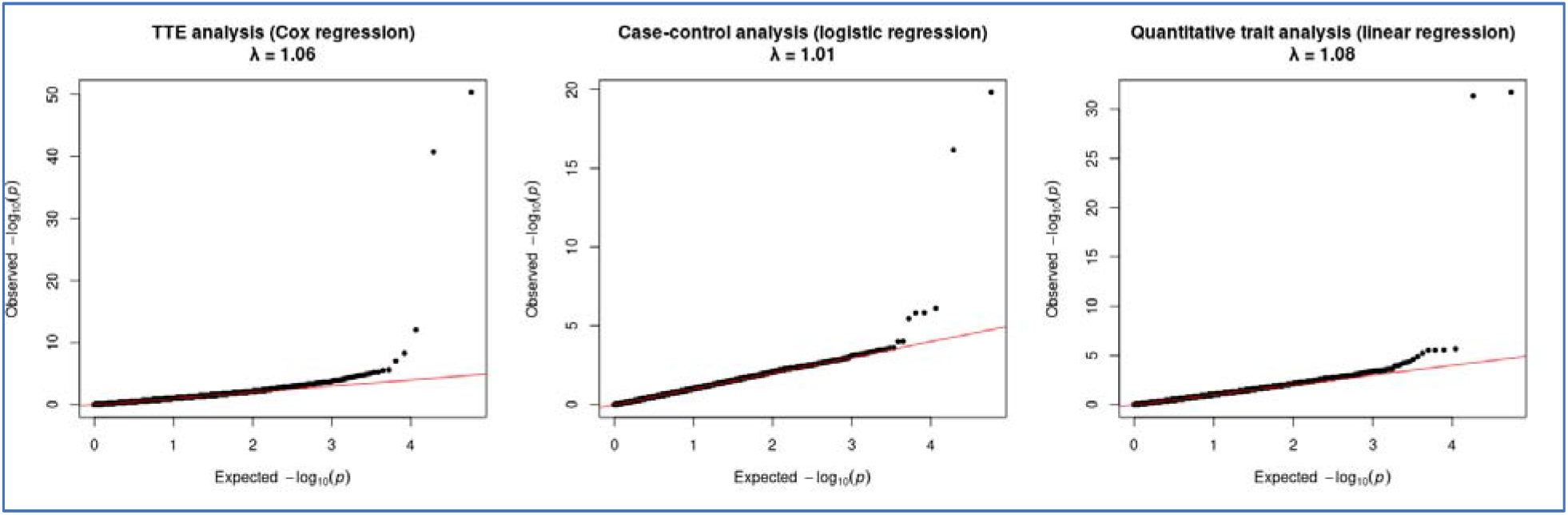
QQ-plots of association p-values from the discovery analysis and corresponding λ_GC_ values quantifying genome-wide inflation.

**Figure S3.**
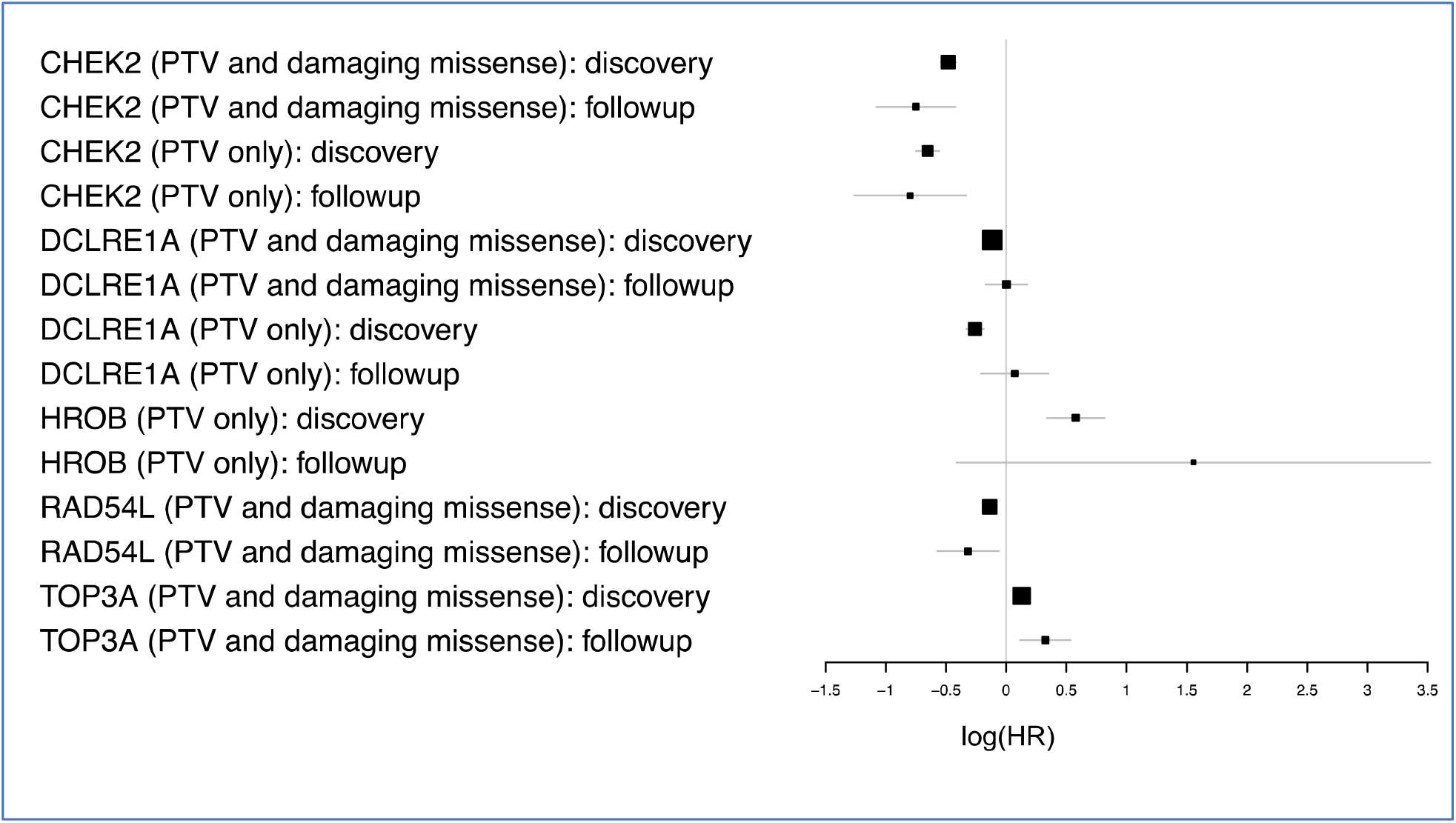
Replication of menopause timing associations in a subset of women who were premenopausal at the initial interview and who underwent follow-up interviews.

**Figure S4.**
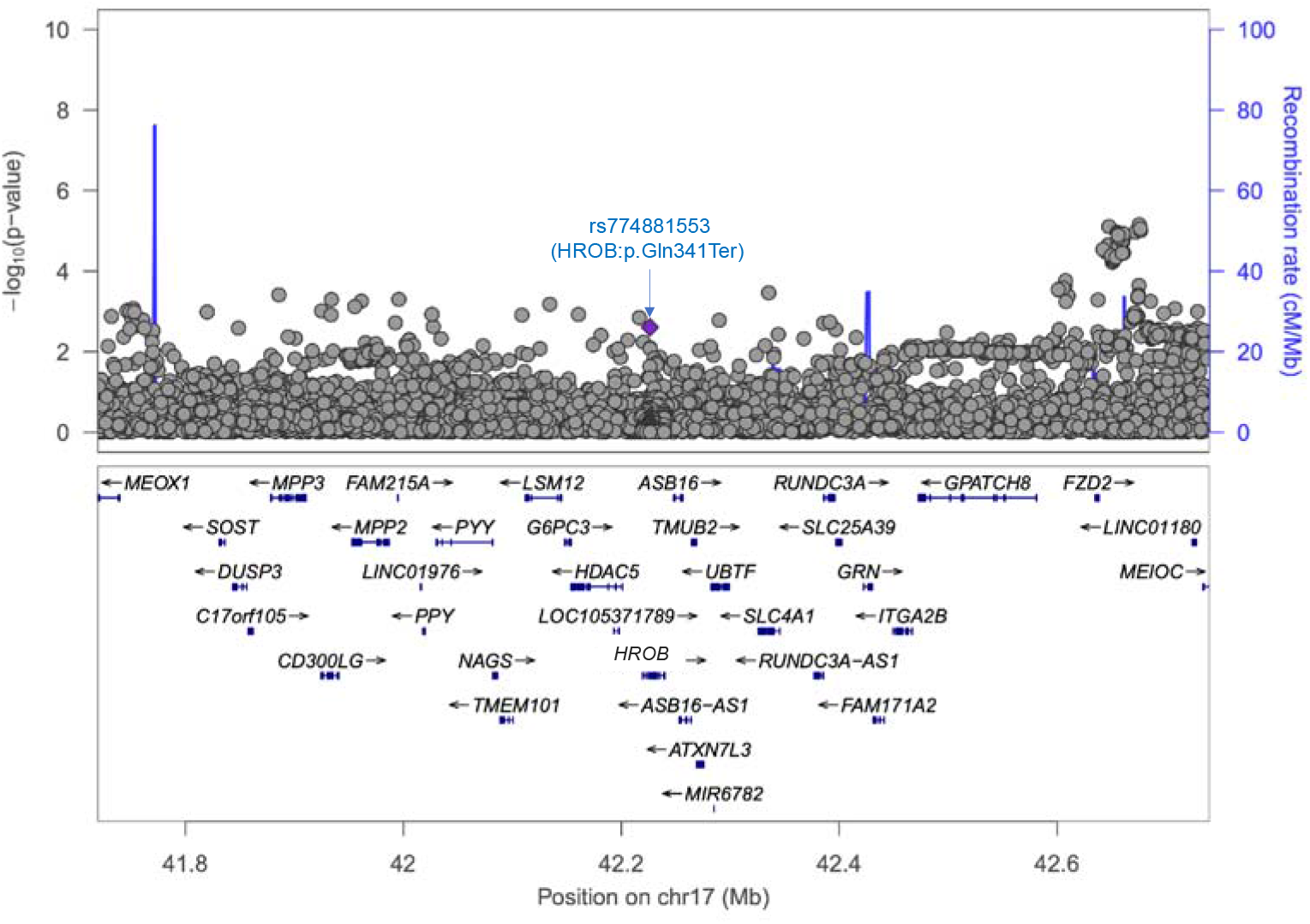
All array and exome single-variant associations with menopause timing at the HROB locus.

**Figure S5.**
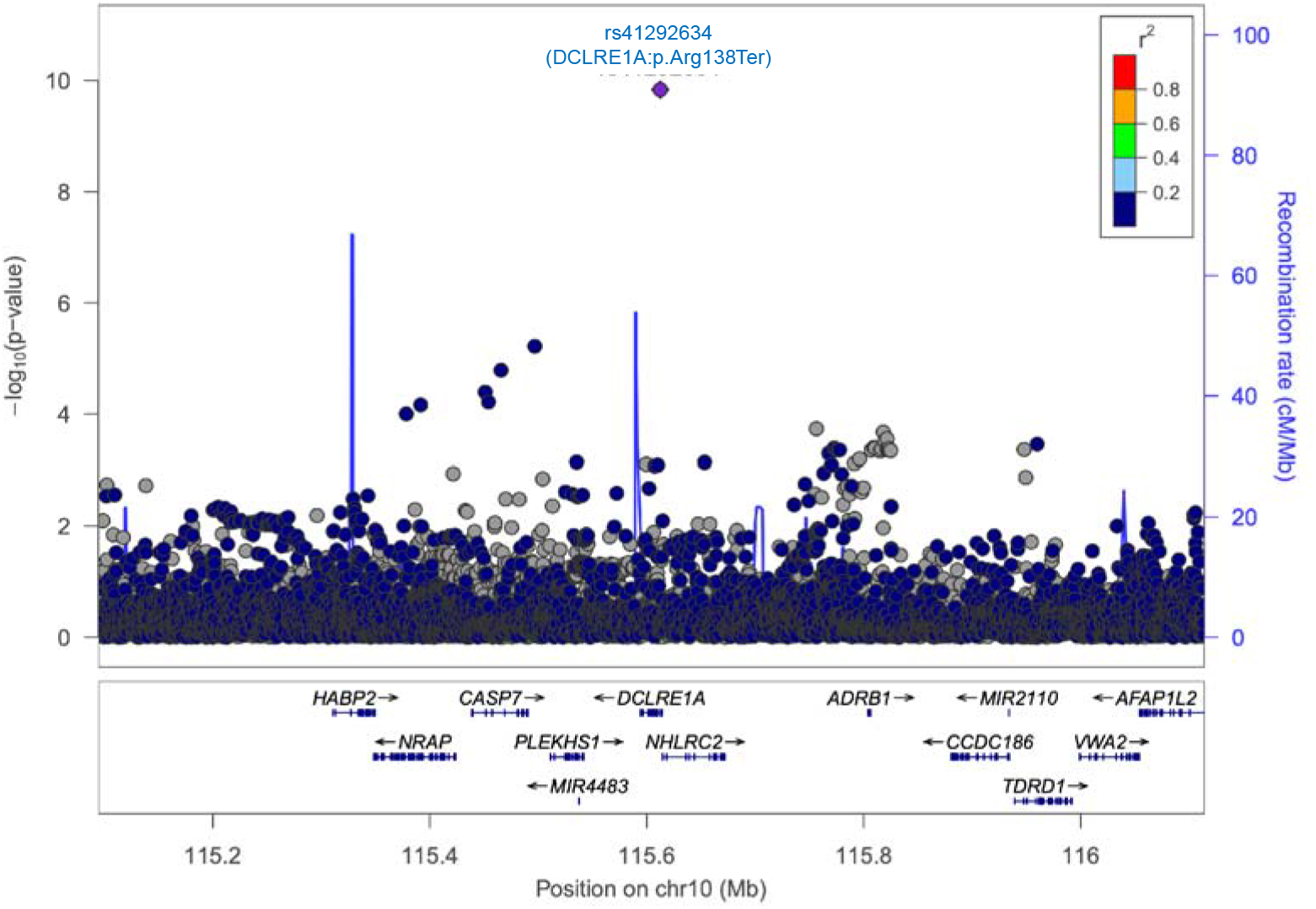
All array and exome single-variant associations with menopause timing at the DCLRE1A locus.

**Figure S6.**
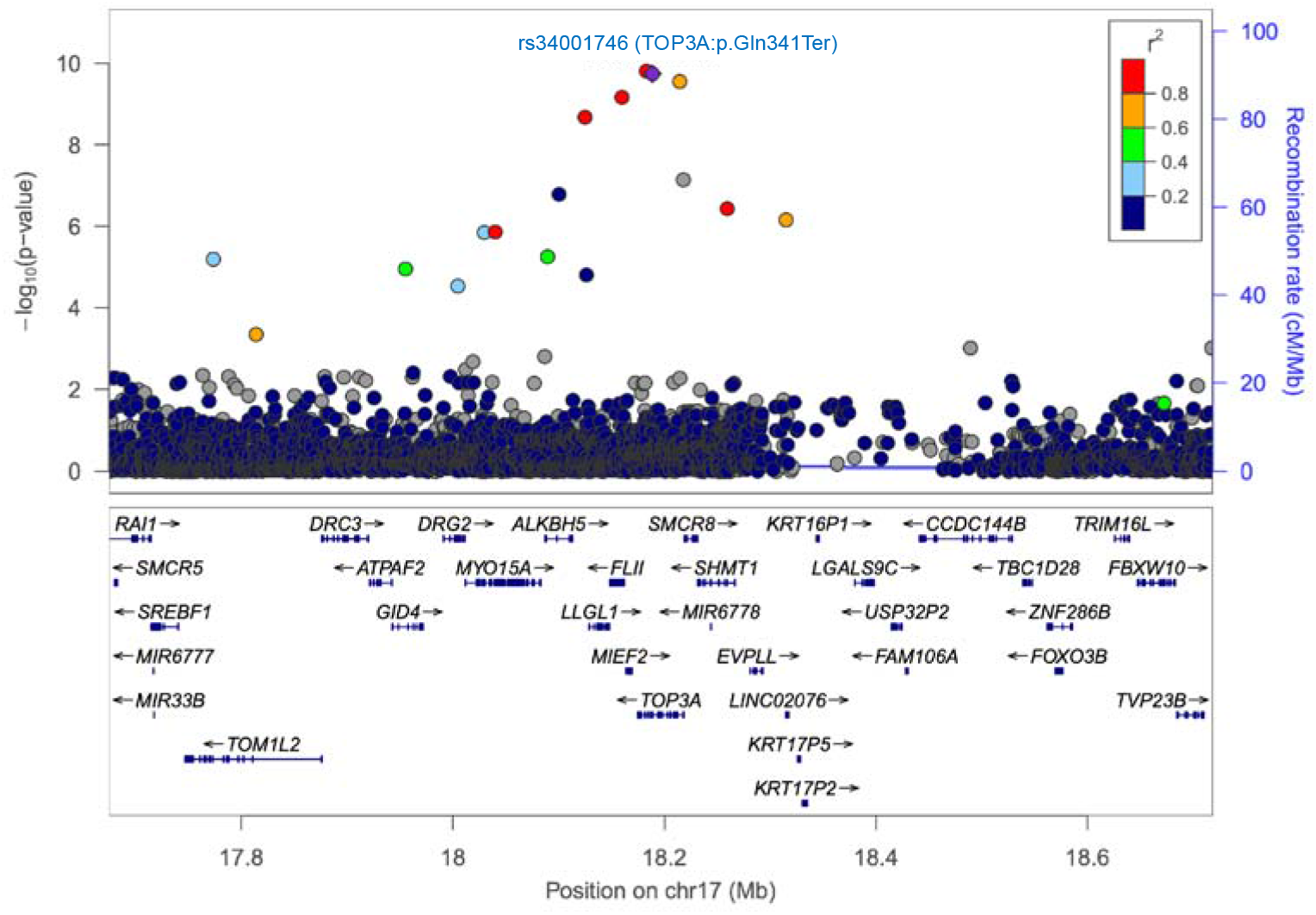
All array and exome single-variant associations with menopause timing at the TOP3A locus.

**Figure S7.**
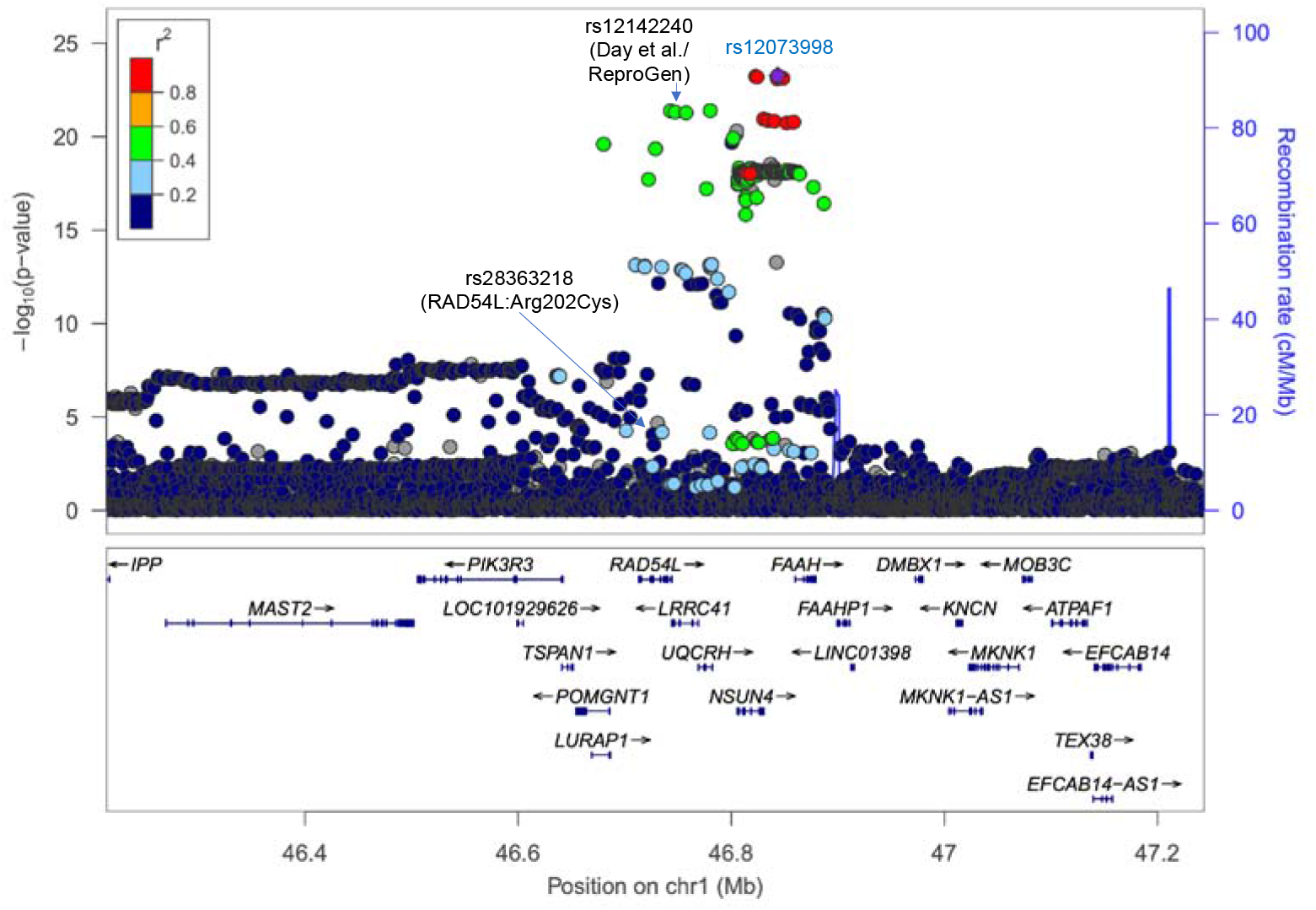
All array and exome single-variant associations with menopause timing at the RAD54L locus.

**Figure S8.**
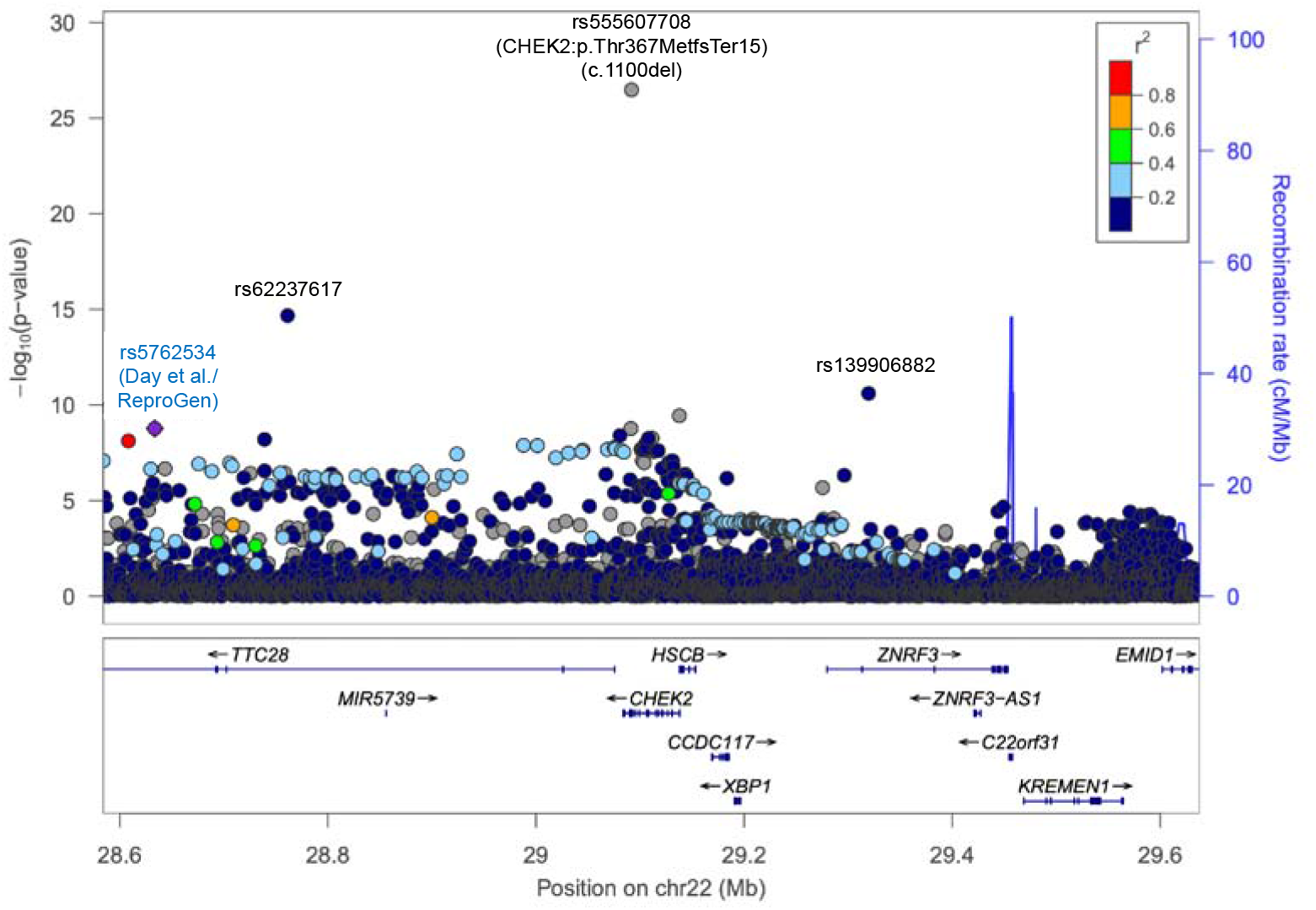
All array and exome single-variant associations with menopause timing at the CHEK2 locus.

**Figure S9.**
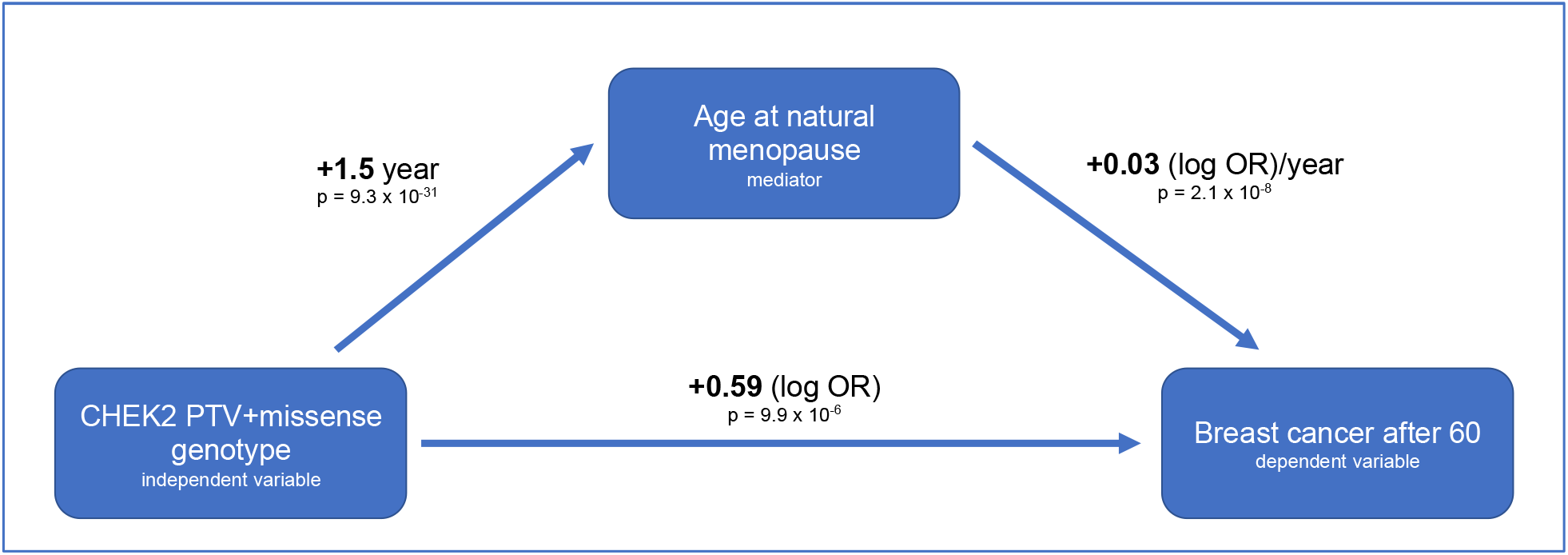
Mediation analysis of the proportion of CHEK2 rare variants (PTVs and damaging missense) breast cancer effect mediated by delaying ANM.

